# Examining side effect variation of antipsychotic treatment in schizophrenia spectrum disorders

**DOI:** 10.1101/2020.07.27.20162727

**Authors:** Maria S. Neumeier, Stephanie Homan, Stefan Vetter, Erich Seifritz, John M. Kane, Maximilian Huhn, Stefan Leucht, Philipp Homan

## Abstract

**Background:** Side effects of antipsychotic drugs play a key role in non-adherence and discontinuation of treatment in schizophrenia spectrum disorders (SSD). Precision medicine aims to minimize such side effects by selecting the right treatment for the right patient. However, to determine the extent of precision medicine that is required, we need to (1) show that there is indeed variation in side effects and (2) estimate the amount of variation in those side effects between patients. While clinical observations suggest that such variation may be considerable, a statistical comparison of side effect variation between active and control treatments is required to confirm this. Here, we hypothesized to find larger side effect variation in treatment compared with control in patients treated with first and second generation antipsychotics.

**Methods:** We included double-blind, placebo-controlled, randomized controlled trials (RCTs) of adults with a diagnosis of SSD and prescription for licensed antipsychotic drugs. Standard deviations of the pre-post treatment differences of weight gain, prolactin levels, and corrected QT (QTc) times were extracted. Data quality and validity were ensured by following the PRISMA guidelines. The outcome measure was the overall variability ratio of treatment to control across RCTs. Individual variability ratios were weighted by the inverse-variance method and entered into a random-effects model.

**Results:** We included N = 16578 patients for weight gain, N = 16633 patients for prolactin levels, and N = 10384 patients for QTc time. Variability ratios (VR) were significantly increased for weight gain (VR = 1.08; 95% CI: 1.02 - 1.14; P = 0.004) and prolactin levels (VR = 1.38; 95% CI: 1.17 - 1.62; P < 0.001) but did not reach significance for QTc time (VR = 1.05; 95% CI: 0.98 - 1.12; P = 0.135).

**Conclusion:** We found increased variability in major side effects in patients with SSD under treatment with second generation antipsychotics, suggesting that subgroups of patients or even individual patients may benefit from improved treatment allocation through stratified or personalized medicine, respectively.

## Introduction

Antipsychotics are a fundamental component in the treatment of schizophrenia spectrum disorders (SSD). Yet, a major problem are side effects which play a key role in nonadherence and discontinuation. ^1–5^ A common hypothesis among researchers and clinicians alike is that although side effects are pervasive, not all patients are equally susceptible.^6^ However, empirical support for this hypothesis is lacking, as randomized controlled trials (RCTs) or conventional meta analyses by design cannot answer whether such side effect variation does exist.^7,8^

To date, studies have established the efficacy, safety, and side effect profiles of antipsychotic medications by averaging these indices across groups of patients. Such studies can provide us with average side effects, but they cannot tell us anything about individual patients or subgroups.^9,10^ Nevertheless, before searching for potential biomarkers that might predict individual susceptibility, we should first quantify the extent to which such predictors are truly needed.

An approach to answering this question is to shift the focus from the means to the variances of side effects.^11^ By comparing the variances between treatment and control groups of RCTs,^12^ greater variability in treatment would indicate that there is a component of variation, the side effect-by-patient or side effect-by-subgroup interaction, that indicates variability of side effects.^11^ Note that this method^13^ has recently been applied for antipsychotics,^7^ antidepressants,^8,14,15^ and brain stimulation.^16^ It is worth noting that these studies found little evidence for treatment effect variation.^7,8,14,15^ Importantly, in the case of pre-post differences used as input for a metaanalysis of variance it is crucial to think carefully about the way the variability ratio is expressed,^12,15,17^ as the use of the coefficient of variation ratio (CVR) that has been proposed as an alternative of the variability ratio (VR)^12^ may lead to unreliable results.^13,17^

A recently published study investigated the individual treatment response in antipsychotics and brought surprising results.^7,18^ By comparing the variability between treatment and control groups, no evidence was found for an increase in variability in the treatment group. What might sound counter-intuitive at first raises the question of how big the need for precision medicine really is. However, that study evaluated the evidence for treatment effect variation. It is possible that although such variation in treatment effects is not as high as sometimes assumed^19^, it does exist in the susceptibility for side effects. In other words, even if there is little variation in response to treatment between patients, there may still be enough variation in side effects to justify a need for precision medicine. If true, then this would support optimization of treatment allocation with respect to side effect profiles.^20^

Side effects that are particularly relevant to antipsychotic treatment include weight gain^5^, hyperprolactinemia, and QTc prolongation.^20^ Weight gain is a frequently observed side effect that can negatively impact one’s physical health and thus may also influence treatment adherence. Every additional kilogram of weight gain can contribute to an increased risk of heart failure,^21^ cardiovascular diesease,^22^ and diabetes.^23^ In addition, treatment discontinuation is often seen in patients with increase of weight under treatment.^24^ High prolactin levels can lead to symptoms like decreased bone mass, gallactorhea, and fertility problems in men and women. Further possible symptoms include menstrual disturbances in female patients and decreased libido and erectile dysfunction in male patients.^25^ These symptoms are frequent, but often under-reported by patients and unnoticed as well as untreated by clinicians.^26, 27^ They furthermore might lead to loss in quality of life and might be a reason for treatment discontinuation^1,28^ and subsequent illness relapse, which together with persistent positive symptoms^29–32^ may severly impact recovery and therapeutic alliance.^33^ Prolongation of QTc was observed in 7 of 14 antipsychotics compared by placebo in the intergroup comparision by Huhn and colleagues.^6^ Importantly, torsade de pointes tachycardia and sudden cardiac death are possible severe consequences of QTc prolongation.^34^ Such cardiac events are one of the factors that lead to the loss of life expectancy observed in patients with SSD.^35–37^

In summary, antipsychotic side effects are highly relevant for long-term outcome and adherence in treatment of positive symptoms in SSD. The question remains whether variability in side effects is high enough to warrant efforts of treatment stratification or personalisation. Here, we compared the variances of side effects including weight gain, prolactin level and QTc-time between treatment and control groups to address this question and to evaluate the evidence for the presence of side effect variability. Based on the clinical impression that patients seem to vary in their susceptibility to side effects, we hypothesized that the variability in side effects would be higher in the treatment compared to the control groups across all published trials of antipsychotics in SSD.^6^

## Methods

### Search strategy and selection criteria

We used the data from the recent meta-analysis by Huhn and colleagues.^6^ That study included placebo-controlled published and unpublished trials investigating orally administered atypical (second generation) antipsychotics and typical (first generation) antipsychotics in adults with schizophrenia spectrum disorders; and excluded patients with first episode psychosis, treatment resistance, mainly negative symptoms, comorbidity with other mental or physical illnesses and relapse-prevention studies. Long- and short-acting intramuscular injections were also excluded (as they are often used in relapse prevention or emergency treatment) and studies from mainland China were excluded because of data quality concerns.^38^ Data sources were MEDLINE, Cochrane Central Register of Controlled Trials (CENTRAL), Embase, Biosis, PsycINFO, PubMed, ClinicalTrials.gov, WHO International Clinical Trials Registry Platform and the US Food and Drug Administration until January 8 2019. Data quality and validity were ensured by following the PRISMA guidelines.^39^ For missing data, we also contacted study authors.

For the analysis, we used the standard deviations of pre-post differences in side effects. The primary outcome was the overall variability ratio of side effects in treatment versus control groups. Standard deviations (SD) and number of patients (N) were extracted for weight gain, prolactin level and QTc time. The units used were kg for weight gain, ng/mL for prolactin levels, and ms for QTc time. Some studies provided data for all of the three side effects, whereas the majority of the studies contained less data (see Results).

### Statistical analysis

If patients or subgroups differ in their susceptibility to side effects, we would expect to observe increased variances in the treatment-compared with the control group. To test this, we computed the log variability ratio (log VR) by comparing the relative variability of side effects under treatment versus control:

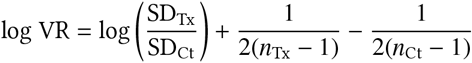

where SD_Tx_ was the reported sample SD for side effects under treatment, SD_Ct_ was the reported sample SD for side effects under control, *n*_Tx_ was the treatment sample size, and *n*_Ct_ the control sample size. The corresponding sampling variance 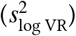 for each comparison can be expressed as follows:

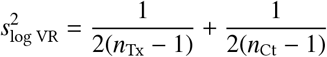

The individual variability ratios were weighted with the inverse of this sampling variance^40^ and entered into a random-effects model to quantify the overall variability ratio of side effects. For better interpretability, results were back-transformed from the logarithmic scale. Here, a variability ratio greater than one would indicate a higher side effect variability in treatment compared to control, whereas a variability ratio smaller than one indicates less side effect variability under treatment compared with control.

### Data and code availability

The analysis was performed from September 2019 to May 2020, using the R package metafor^40^(version 2.1.0). The manuscript was produced with the R packages rmarkdown (version 2.1); represearch (version 0.0.0.9000; https://github.com/phoman/represearch/); knitr (version 1.26); and papaja (version 0.1.0.9942). All data and code are freely available online to ensure reproducibility at https://github.com/homanlab/sideeffects/.

## Results

### Overall reporting details

Together, we screened N = 151 studies from the original metaanalysis by Huhn and colleagues^6^ as these studies reported data on at least one of the three side effects that we were interested in. Of these studies, N = 94 (62%) had missing variance measures despite reported means for at least one of the three side effects. We thus included the N = 60 (40%) studies that did report variance measures for at least one of the side effects of interest.

### Weight gain

For weight gain, we included 51 RCTs, yielding 72 comparisons of antipsychotic drugs with placebo to investigate the individual occurrence of weight gain in patients. All together we included N = 16578 patients diagnosed with schizophrenia or schizoaffective disorder. There were 11373 (69%) patients randomly allocated to the treatment group, and 5205 (31%) to the placebo group. Overall, the variability for weight gain was higher under treatment than under control (VR = 1.08; 95% CI: 1.02 - 1.14; P = 0.004; Figure 1). Individual comparisons between drugs across studies indicated marked differences between individual antipsychotics (VR = 1.08; 95% CI: 1.02 - 1.14; P = 0.004; Figure 2).

**Figure 1.**
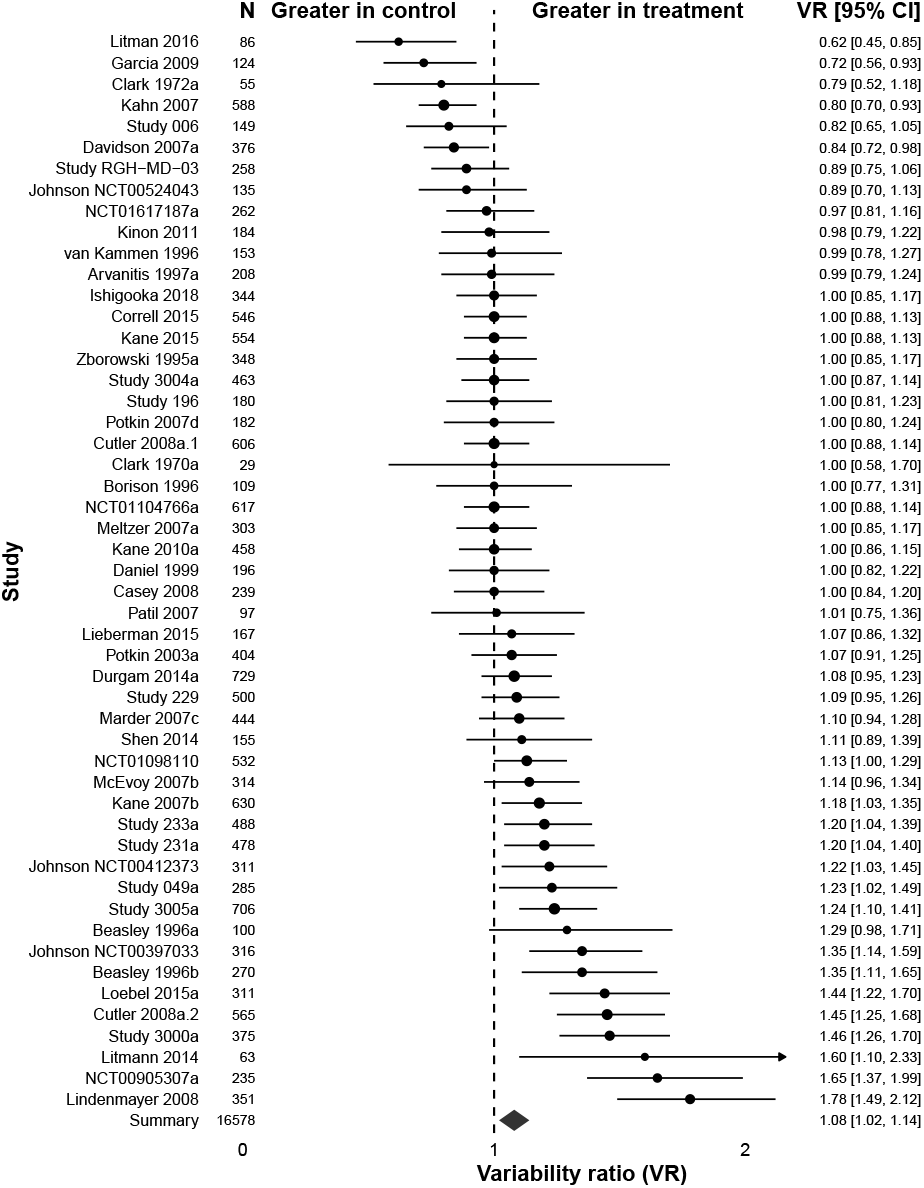
Variability ratio for weight gain. The forest plot shows the VR together with its 95% confidence interval (CI) for treatment versus control. All included studies^41–93^ are also listed in Table S1.

**Figure 2.**
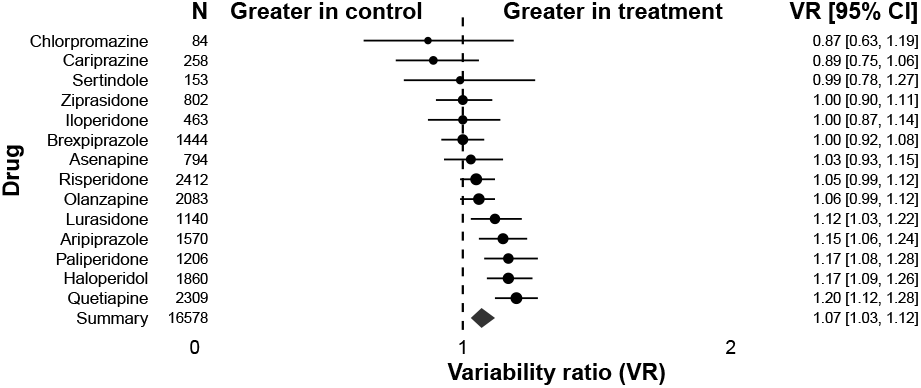
Variability ratio for weight gain for individual antipsychotics. The forest plot shows the VR together with its 95% confidence interval (CI) for treatment versus control. All included studies^41–93^ are also listed in Table S1.

### Hyperprolactinemia

For hyperprolactinemia, we included 50 RCTs, with 71 comparisons of antipsychotic drugs with placebo. All together we included N = 16633 patients diagnosed with schizophrenia or schizoaffective disorder. There were 11409 (69%) patients randomly allocated to the treatment group, and 5224 (31%) to the placebo group. Overall, the variability for hyperprolactinemia was higher under treatment than under control (VR = 1.38; 95% CI: 1.17 - 1.62; P < 0.001; Figure 3). Individual comparisons between drugs across studies indicated marked differences between individual antipsychotics (VR = 1.38; 95% CI: 1.17 - 1.62; P < 0.001; Figure 4).

**Figure 3.**
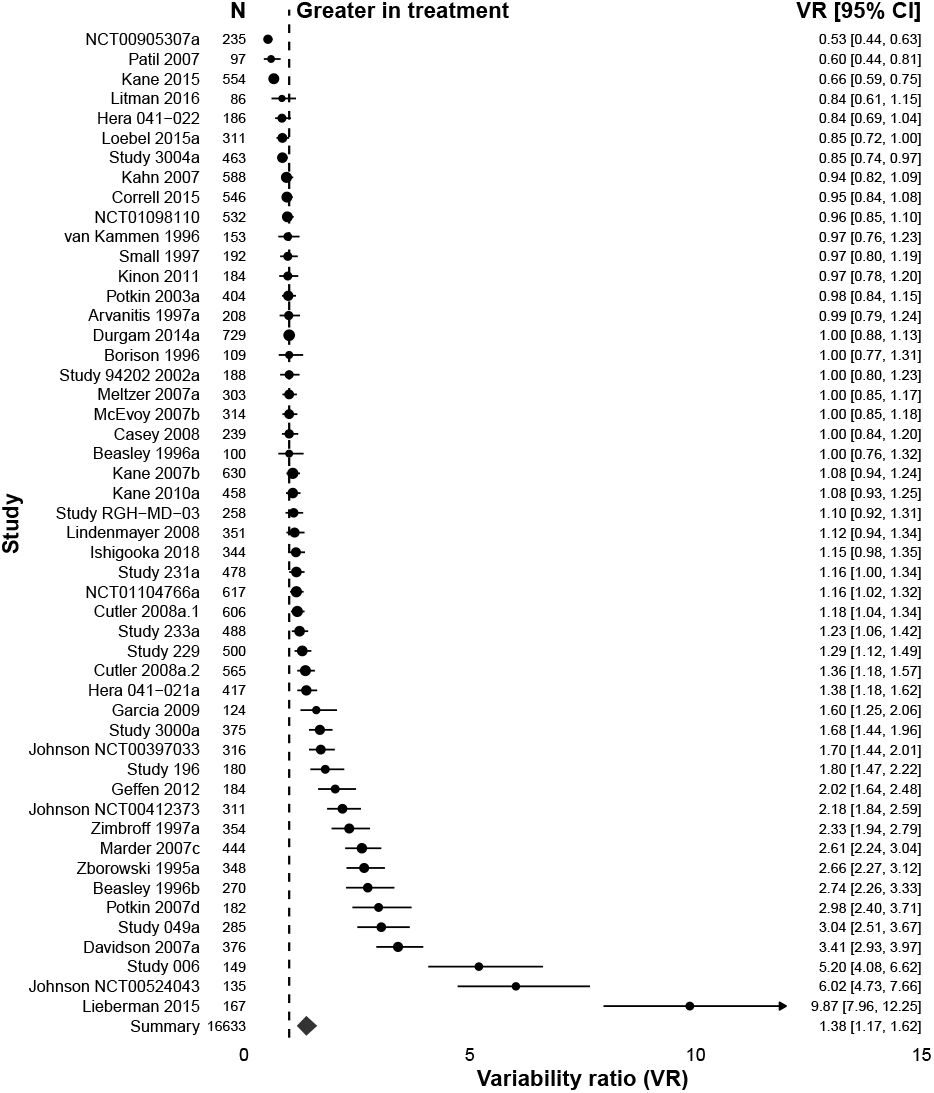
Variability ratio for hyperprolactinemia. The forest plot shows the VR together with its 95% confidence interval (CI) for treatment versus control. All included studies^41,42,44–48,50–58,60,62,64,65,67–69,72,74–79,82–90,92–100^ are also listed in Table S1.

**Figure 4.**
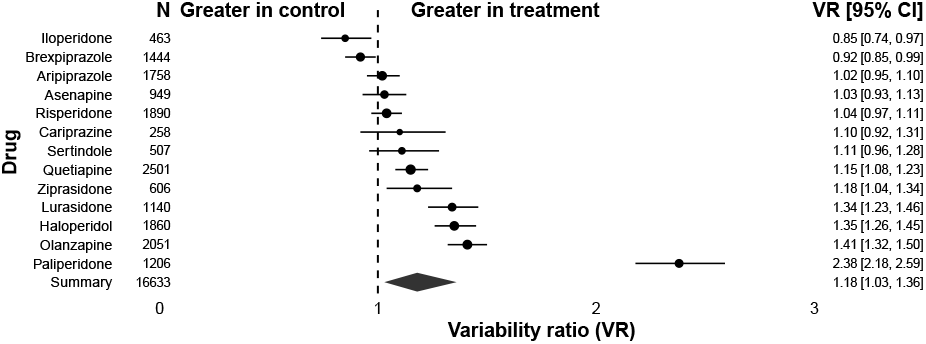
Variability ratio for hyperprolactinemia for individual antipsychotics. The forest plot shows the VR together with its 95% confidence interval (CI) for treatment versus control. All included studies^41,42,44–48,50–58,60,62,64,65,67–69,72,74–79,82–90,92–100^ are also listed in Table S1.

### QTc prolongation

For QTc prolongation, we included 29 RCTs, with 46 comparisons of antipsychotic drugs with placebo. All together we included N = 10384 patients diagnosed with schizophrenia or schizoaffective disorder. There were 7439 (72%) patients randomly allocated to the treatment group, and 2945 (28.00%) to the placebo group. Even though the variability for QTc prolongation was higher under treatment than under control, the difference did not reach statistical significance (VR = 1.05; 95% CI: 0.98 - 1.12; P = 0.135; Figure 5).

**Figure 5.**
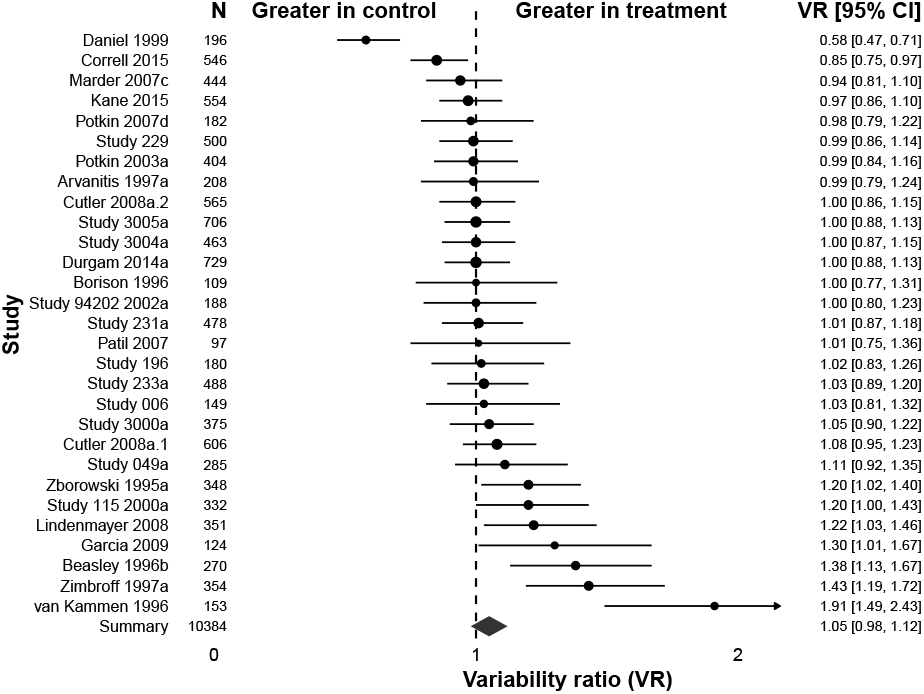
Variability ratio for QTc prolongation. The forest plot shows the VR together with its 95% confidence interval (CI) for treatment versus control. All included studies^42,45,51,52,54–60,62,68,70,74,76–80,85,86,89,93,94,99,100^ are also listed in Table S1.

However, individual comparisons between drugs across studies indicated marked differences between individual antipsychotics, with sertindole and haloperidol leading to significant increases in variability compared to control (VR = 1.05; 95% CI: 0.98 - 1.12; P = 0.135; Figure 6).

**Figure 6.**
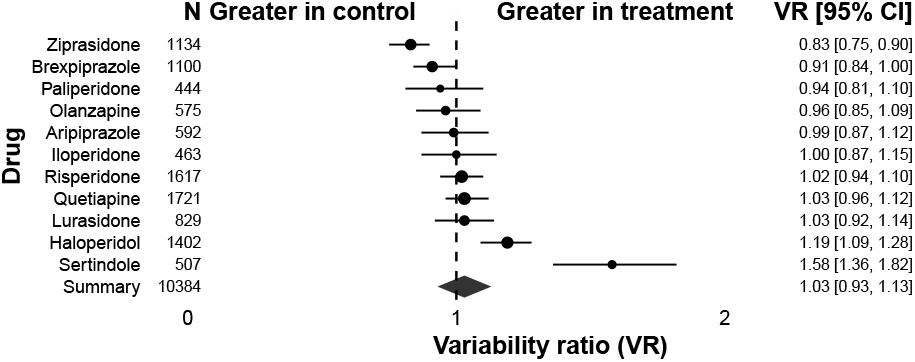
Variability ratio for QTC prolongation for individual antipsychotics. The forest plot shows the VR together with its 95% confidence interval (CI) for treatment versus control. All included studies^42,45,51,52,54–60,62,68,70,74,76–80,85,86,89,93,94,99,100^ are also listed in Table S1.

## Discussion

### Summary

This study assessed the variability in the three major side effects of antipsychotic treatment in schizophrenia spectrum disorders. We focused on side effects because their occurrence has a great impact on treatment adherence and pyhsical health of patients, and clinical experience suggests a potential to improve treatment allocation by taking into account the variability in side effect occurrence. We wanted to quantify the evidence in support of this experience, relevant for clinicians as much as for translational researchers. We also know from clinical trials and meta-analyses that some antipsychotics are more associated with specific side effects than others. For example, clozapine and olanzapine are strongly associated with weight gain,^6,20,101^ QTc-time prolongation is most distinct in sertindole and amisulpride^6^, and prolactin level elevation in paliperidone and risperidone.^6^ However, these data cannot address the question whether there is variability in subgroups or individual patients. Such side effect-by-subgroup or side effect-by-patient interaction would be a prime example for the need of a more stratified or personalized medicine, respectively, which allocates treatments according to side effect profiles of subgroups or individual patients. The presence of such subgroups or individual patients would result in an increase of the side effect variability of treated patients compared to those who received placebo.^7,13^. The amount of this increase can be captured by the variability ratio (VR) which compares the variability of treatment versus control for each side effect. Evaluating all studies that reported variance measures for at least one of the investigated side effects,^6^ we found that the reporting of standard deviations was often incomplete. In terms of variability of side effects, we found that the variability for weight gain and prolactin elevation was indeed significantly increased in patients who received treatment compared to those who received placebo. For QTc prolongation, this increase was did not reach significance. Together, our results suggest that there is indeed marked variability in the occurrence of side effects in antipsychotic treatment.

### Reporting

Altogether we included 43595 patients from 60 studies. Only for about 40% of studies included in a previous meta-analysis^6^ variance data for at least one of the side effects of interest (weight gain, prolactin levels, QTc prolongation) were available. In about 62% of the studies included^6^ incomplete data existed such that means were reported without a measure of variance. Although we did contact authors for missing data whenever possible, we received missing data only for three studies. In summary, consistent reporting of antipsychotic side effects, specifically with respect to variability measures, is currently missing in the literature and should be improved in future studies.

### Weight gain

Weight gain in antipsychotics, especially in second generation antipsychotics,^102^ is a severe side effect that can contribute to metabolic dysregulation. Importantly, every kg of weight gain leads to a linear increase in the risk of cardiovascular diseases^22^, heart failure^21^, and diabetes^23^. Clozapine, olanzapine, zotepine and sertindole have the most severe impact in gaining weight. Some studies showed that a lower BMI at baseline^103^ and sex^104^ can lead to more weight gain, whereas other studies found that male sex and higher BMI at baseline are related to a higher risk of metabolic distrubances.^20^ Our findings provide evidence that some patients are indeed more susceptible to antipsychotic weight gain than others. As antipsychotics in the treatment for schizophrenia and related diseases is often recommended to be taken as a relapse prevention for a longer period,^105,106^ patients are likely to gain more weight during their treatment over months and years. Together, this suggests that there is a potential to improve long-term health and adherence by identifying the subgroups or individual patients that are particularly prone to weight gain. Preliminary evidence suggests that a dysregulated striatal reward circuit contributes to weight gain susceptbility.^5,107^

### Hyperprolactinemia

Prolactin level elevations occur in up to 70% of patients^108^ under the treatment with antipsychotic drugs. By blocking dopamine D2 receptors on lacotroph cells a disinhibition of the synthesis and secretion of prolactin is observed.^109,110^ This can lead to both, shortand long-term side effects with potentially severe impact on our patients health. Typical short time effects include galactorrhea, gynecomastia, menstrual irregularities, and sexual dysfunction; a typical long-term result is osteoporosis.^111,112^ and a potentially increased risk in developing breast cancer in association with hyperprolactinemia.^113,114^ Our findings suggest that these risks may be particularly relevant for some patients but not other patients. For example, a previous study found that prolactin level elevations are more pronounced and more frequent in women than in men.^115^ In addition, some antipsychotics such as amisulprid, risperidone, and paliperidone are linked to a greater elevation of prolactin.^6,115^ In summary, and in line with the weight gain findings, this suggests that there is a potential to improve long-term health and antipsychotic adherence by identifying the subgroups or individual patients that are particularly likely to develop prolactine elevations under antipsychotic treatment.

### QTc prolongation

Prolongation of QTc is another important antipsychotic side effect as cardiovascular diseases remain the most common cause of natural mortality in schizophrenia spectrum disorders.^116^ Users of antipsychotic medication are reported to have higher rates of sudden cardiac death than nonusers.^117^ Prolongation of QTc (longer than 450 ms in men and longer than 470 ms in women, respectively, when corrected with Bazetts Formula^118^) can contribute to this.^34^ A prolongation of QTc can lead to torsade de pointes and subsequently to sudden death.^119,120^ The molecular pathway of this side effect is not completely understood.^121^ It is known, however, that some medications such as sertindole, amisulprid, ziprasidone lead to more QTc prolongation than others.^6^ Our findings suggest that although QTc prolongation varies between subgroups or patients this increased variability is not statistically significant, potentially because of a smaller number of studies available which decreased the statistical power. Previous studies suggest that risk factors may include female sex, comorbid cardiovascular disease, high drug dosages, and electrolyte disturbances.^122^

### Limitations and strengths

Our meta-analysis had some limitations. First, the occurrence of side effects might be a dosage dependent effect, which could reflect a higher/different VR in some studies. Second, the level of prolactin can be highly variable based on multiple biological and methodological factors such as stress, diurnal variation and type of assay performed. This might explain the surprising difference in prolactin level variability between risperidone and paliperidone, two highly similar drugs. Third, for QTc, a reduced number of studies was available, potentially reducing statistical power to detect a significant variability increase. Finally, our method cannot determine whether the increased variability is due to variability differences in subgroups or individual patients.^11^. The particular strength of our study is that we included all available studies of antipsychotic treatment in SSD reporting variability measures for side effects of interest. To our knowledge, this is the first comprehensive study that provides evidence for substantial variability in side effects.

### Conclusion

Our findings suggest that there is enough variability in two major side effects (weight gain and prolactin elevation) to assume that subgroups of patients or even individual patients may benefit from improved treatment allocation through stratified or personalized medicine, respectively. Such efforts in precision medicine might be crucial to improve adherence^123^ and long-term health under antipsychotic treatment.

## Data Availability

All data and code are freely available online to ensure reproducibility at https://github.com/homanlab/sideeffects/.

## Acknowledgements

The authors thank Majnu John, PhD, for advice on the analysis of the current study and Ellen Ji, PhD, for her thoughtful comments on the manuscript. These individuals received no additional compensation, outside of their usual salary, for their contributions.

## Supplementary Information

### Supplementary Tables

**Table S1.** All study arms with references

| Study Arm | Year | Sample Size | Drug | Weight | Prolactin | QTc |
| --- | --- | --- | --- | --- | --- | --- |
| Ishigooka 2018<sup>53</sup> | 2018 | 116 | Placebo | Yes | Yes | No |
| Ishigooka 2018<sup>53</sup> | 2018 | 228 | Brexpiprazole | Yes | Yes | No |
| Litman 2016<sup>41</sup> | 2016 | 55 | Placebo | Yes | Yes | No |
| Litman 2016<sup>41</sup> | 2016 | 31 | Risperidone | Yes | Yes | No |
| NCT01104766a<sup>64</sup> | 2015 | 153 | Placebo | Yes | Yes | No |
| NCT01104766a<sup>64</sup> | 2015 | 152 | Aripiprazole | Yes | Yes | No |
| NCT01104766b<sup>64</sup> | 2015 | 312 | Cariprazine | Yes | Yes | No |
| Lieberman 2015<sup>75</sup> | 2015 | 85 | Placebo | Yes | Yes | No |
| Lieberman 2015<sup>75</sup> | 2015 | 82 | Risperidone | Yes | Yes | No |
| Kane 2015<sup>124</sup> | 2015 | 184 | Placebo | Yes | Yes | Yes |
| Kane 2015<sup>124</sup> | 2015 | 370 | Brexpiprazole | Yes | Yes | Yes |
| Correll 2015<sup>54</sup> | 2015 | 184 | Placebo | Yes | Yes | Yes |
| Correll 2015<sup>54</sup> | 2015 | 362 | Brexpiprazole | Yes | Yes | Yes |
| NCT01098110<sup>82</sup> | 2015 | 174 | Placebo | Yes | Yes | No |
| NCT01098110<sup>82</sup> | 2015 | 358 | Asenapine | Yes | Yes | No |
| NCT01617187a<sup>49</sup> | 2015 | 113 | Asenapine | Yes | No | No |
| NCT01617187a<sup>49</sup> | 2015 | 103 | Placebo | Yes | No | No |
| NCT01617187b<sup>49</sup> | 2015 | 46 | Olanzapine | Yes | No | No |
| NCT00905307a<sup>92</sup> | 2015 | 50 | Aripiprazole | Yes | Yes | No |
| NCT00905307a<sup>92</sup> | 2015 | 95 | Placebo | Yes | Yes | No |
| NCT00905307b<sup>92</sup> | 2015 | 90 | Brexpiprazole | Yes | Yes | No |
| Loebel 2015a<sup>90</sup> | 2015 | 199 | Lurasidone | Yes | Yes | No |
| Loebel 2015a<sup>90</sup> | 2015 | 112 | Placebo | Yes | Yes | No |
| Litmann 2014<sup>91</sup> | 2014 | 41 | Placebo | Yes | No | No |
| Litmann 2014<sup>91</sup> | 2014 | 22 | Olanzapine | Yes | No | No |
| Schmidt 2014<sup>96</sup> | 2014 | 93 | Olanzapine | Yes | Yes | No |
| Shen 2014<sup>81</sup> | 2014 | 78 | Placebo | Yes | No | No |
| Shen 2014<sup>81</sup> | 2014 | 77 | Olanzapine | Yes | No | No |
| Durgam 2014a<sup>77</sup> | 2014 | 151 | Placebo | Yes | Yes | Yes |
| Durgam 2014a<sup>77</sup> | 2014 | 140 | Risperidone | Yes | Yes | Yes |
| Durgam 2014b<sup>77</sup> | 2014 | 438 | Cariprazine | Yes | Yes | Yes |
| Geffen 2012<sup>97</sup> | 2012 | 91 | Risperidone | No | Yes | No |
| Geffen 2012<sup>97</sup> | 2012 | 93 | Placebo | No | Yes | No |
| Kinon 2011<sup>50</sup> | 2011 | 62 | Olanzapine | Yes | Yes | No |
| Kinon 2011<sup>50</sup> | 2011 | 122 | Placebo | Yes | Yes | No |
| Kane 2010a<sup>67</sup> | 2010 | 115 | Haloperidol | Yes | Yes | No |
| Kane 2010a<sup>67</sup> | 2010 | 123 | Placebo | Yes | Yes | No |
| Kane 2010b<sup>67</sup> | 2010 | 220 | Asenapine | Yes | Yes | No |
| Study 006<sup>45</sup> | 2010 | 99 | Lurasidone | Yes | Yes | Yes |
| Study 006<sup>45</sup> | 2010 | 50 | Placebo | Yes | Yes | Yes |
| Study 049a<sup>100</sup> | 2010 | 73 | Haloperidol | Yes | Yes | Yes |
| Study 049a<sup>100</sup> | 2010 | 72 | Placebo | Yes | Yes | Yes |
| Study 049b<sup>100</sup> | 2010 | 140 | Lurasidone | Yes | Yes | Yes |
| Study 196<sup>57</sup> | 2010 | 90 | Placebo | Yes | Yes | Yes |
| Study 196<sup>57</sup> | 2010 | 90 | Lurasidone | Yes | Yes | Yes |
| Study 229<sup>78</sup> | 2010 | 372 | Lurasidone | Yes | Yes | Yes |
| Study 229<sup>78</sup> | 2010 | 128 | Placebo | Yes | Yes | Yes |
| Study 231a<sup>86</sup> | 2010 | 116 | Placebo | Yes | Yes | Yes |
| Study 231a<sup>86</sup> | 2010 | 123 | Olanzapine | Yes | Yes | Yes |
| Study 231b<sup>86</sup> | 2010 | 239 | Lurasidone | Yes | Yes | Yes |
| Study 233a<sup>85</sup> | 2010 | 122 | Placebo | Yes | Yes | Yes |
| Study 233a<sup>85</sup> | 2010 | 120 | Quetiapine | Yes | Yes | Yes |
| Study 233b<sup>85</sup> | 2010 | 246 | Lurasidone | Yes | Yes | Yes |
| Garcia 2009<sup>42</sup> | 2009 | 60 | Haloperidol | Yes | Yes | Yes |
| Garcia 2009<sup>42</sup> | 2009 | 64 | Placebo | Yes | Yes | Yes |
| Hera 041-021a<sup>125</sup> | 2009 | 208 | Asenapine | No | Yes | No |
| Hera 041-021a<sup>125</sup> | 2009 | 106 | Placebo | No | Yes | No |
| Hera 041-021b<sup>125</sup> | 2009 | 103 | Olanzapine | No | Yes | No |
| Hera 041-022<sup>126</sup> | 2009 | 93 | Olanzapine | No | Yes | No |
| Hera 041-022<sup>126</sup> | 2009 | 93 | Placebo | No | Yes | No |
| Casey 2008<sup>72</sup> | 2008 | 120 | Risperidone | Yes | Yes | No |
| Casey 2008<sup>72</sup> | 2008 | 119 | Placebo | Yes | Yes | No |
| Cutler 2008a<sup>60</sup> | 2008 | 151 | Ziprasidone | Yes | Yes | Yes |
| Cutler 2008a<sup>60</sup> | 2008 | 152 | Placebo | Yes | Yes | Yes |
| Cutler 2008b<sup>60</sup> | 2008 | 303 | Iloperidone | Yes | Yes | Yes |
| Johnson NCT00397033<sup>88</sup> | 2008 | 209 | Paliperidone | Yes | Yes | No |
| Johnson NCT00397033<sup>88</sup> | 2008 | 107 | Placebo | Yes | Yes | No |
| Johnson NCT00412373<sup>98</sup> | 2008 | 95 | Placebo | Yes | Yes | No |
| Johnson NCT00412373<sup>98</sup> | 2008 | 216 | Paliperidone | Yes | Yes | No |
| Johnson NCT00524043<sup>48</sup> | 2008 | 70 | Paliperidone | Yes | Yes | No |
| Johnson NCT00524043<sup>48</sup> | 2008 | 65 | Placebo | Yes | Yes | No |
| Lindenmayer 2008<sup>93</sup> | 2008 | 267 | Quetiapine | Yes | Yes | Yes |
| Lindenmayer 2008<sup>93</sup> | 2008 | 84 | Placebo | Yes | Yes | Yes |
| Study 3000a<sup>56</sup> | 2008 | 127 | Placebo | Yes | Yes | Yes |
| Study 3000a<sup>56</sup> | 2008 | 124 | Haloperidol | Yes | Yes | Yes |
| Study 3000b<sup>56</sup> | 2008 | 124 | Iloperidone | Yes | Yes | Yes |
| Study 3004a<sup>56</sup> | 2008 | 156 | Placebo | Yes | Yes | Yes |
| Study 3004a<sup>56</sup> | 2008 | 154 | Iloperidone | Yes | Yes | Yes |
| Study 3004b<sup>56</sup> | 2008 | 153 | Risperidone | Yes | Yes | Yes |
| Study 3005a<sup>56</sup> | 2008 | 157 | Risperidone | Yes | No | Yes |
| Study 3005a<sup>56</sup> | 2008 | 160 | Placebo | Yes | No | Yes |
| Study 3005b<sup>56</sup> | 2008 | 389 | Iloperidone | Yes | No | Yes |
| Study RGH-MD-03<sup>47</sup> | 2008 | 130 | Placebo | Yes | Yes | No |
| Study RGH-MD-03<sup>47</sup> | 2008 | 128 | Cariprazine | Yes | Yes | No |
| Cutler 2008a<sup>60</sup> | 2008 | 117 | Placebo | Yes | Yes | Yes |
| Cutler 2008a<sup>60</sup> | 2008 | 448 | Quetiapine | Yes | Yes | Yes |
| Davidson 2007a<sup>46</sup> | 2007 | 123 | Placebo | Yes | Yes | No |
| Davidson 2007a<sup>46</sup> | 2007 | 128 | Olanzapine | Yes | Yes | No |
| Davidson 2007b<sup>46</sup> | 2007 | 125 | Paliperidone | Yes | Yes | No |
| Kahn 2007<sup>44</sup> | 2007 | 118 | Placebo | Yes | Yes | No |
| Kahn 2007<sup>44</sup> | 2007 | 470 | Quetiapine | Yes | Yes | No |
| McEvoy 2007b<sup>83</sup> | 2007 | 206 | Aripiprazole | Yes | Yes | No |
| McEvoy 2007b<sup>83</sup> | 2007 | 108 | Placebo | Yes | Yes | No |
| Meltzer 2007a<sup>65</sup> | 2007 | 149 | Placebo | Yes | Yes | No |
| Meltzer 2007a<sup>65</sup> | 2007 | 154 | Risperidone | Yes | Yes | No |
| Kane 2007b<sup>84</sup> | 2007 | 127 | Placebo | Yes | Yes | No |
| Kane 2007b<sup>84</sup> | 2007 | 128 | Olanzapine | Yes | Yes | No |
| Kane 2007c<sup>84</sup> | 2007 | 375 | Paliperidone | Yes | Yes | No |
| Marder 2007c<sup>79</sup> | 2007 | 110 | Placebo | Yes | Yes | Yes |
| Marder 2007c<sup>79</sup> | 2007 | 224 | Paliperidone | Yes | Yes | Yes |
| Marder 2007d<sup>79</sup> | 2007 | 110 | Olanzapine | Yes | Yes | Yes |
| Patil 2007<sup>74</sup> | 2007 | 34 | Olanzapine | Yes | Yes | Yes |
| Patil 2007<sup>74</sup> | 2007 | 63 | Placebo | Yes | Yes | Yes |
| Potkin 2007d<sup>58</sup> | 2007 | 62 | Placebo | Yes | Yes | Yes |
| Potkin 2007d<sup>58</sup> | 2007 | 60 | Risperidone | Yes | Yes | Yes |
| Potkin 2007c<sup>58</sup> | 2007 | 60 | Asenapine | Yes | Yes | Yes |
| Potkin 2003a<sup>76</sup> | 2003 | 202 | Aripiprazole | Yes | Yes | Yes |
| Potkin 2003a<sup>76</sup> | 2003 | 103 | Placebo | Yes | Yes | Yes |
| Potkin 2003b<sup>76</sup> | 2003 | 99 | Risperidone | Yes | Yes | Yes |
| Kane 2002b<sup>68</sup> | 2002 | 104 | Haloperidol | Yes | Yes | Yes |
| Study 94202 2002a<sup>127</sup> | 2002 | 61 | Aripiprazole | No | Yes | Yes |
| Study 94202 2002a<sup>127</sup> | 2002 | 64 | Placebo | No | Yes | Yes |
| Study 94202 2002b<sup>127</sup> | 2002 | 63 | Haloperidol | No | Yes | Yes |
| Study 115 2000a<sup>128</sup> | 2000 | 83 | Placebo | No | No | Yes |
| Study 115 2000a<sup>128</sup> | 2000 | 164 | Ziprasidone | No | No | Yes |
| Study 115 2000b<sup>128</sup> | 2000 | 85 | Haloperidol | No | No | Yes |
| Daniel 1999<sup>70</sup> | 1999 | 92 | Placebo | Yes | No | Yes |
| Daniel 1999<sup>70</sup> | 1999 | 104 | Ziprasidone | Yes | No | Yes |
| Arvanitis 1997a<sup>52</sup> | 1997 | 51 | Placebo | Yes | Yes | Yes |
| Arvanitis 1997a<sup>52</sup> | 1997 | 105 | Quetiapine | Yes | Yes | Yes |
| Arvanitis 1997b<sup>52</sup> | 1997 | 52 | Haloperidol | Yes | Yes | Yes |
| Small 1997<sup>95</sup> | 1997 | 96 | Quetiapine | No | Yes | No |
| Small 1997<sup>95</sup> | 1997 | 96 | Placebo | No | Yes | No |
| Zimbroff 1997a<sup>99</sup> | 1997 | 144 | Sertindole | No | Yes | Yes |
| Zimbroff 1997a<sup>99</sup> | 1997 | 73 | Placebo | No | Yes | Yes |
| Zimbroff 1997b<sup>99</sup> | 1997 | 137 | Haloperidol | No | Yes | Yes |
| Beasley 1996a<sup>87</sup> | 1996 | 50 | Placebo | Yes | Yes | No |
| Beasley 1996a<sup>87</sup> | 1996 | 50 | Olanzapine | Yes | Yes | No |
| Beasley 1996b<sup>89</sup> | 1996 | 69 | Haloperidol | Yes | Yes | Yes |
| Beasley 1996b<sup>89</sup> | 1996 | 68 | Placebo | Yes | Yes | Yes |
| Beasley 1996c<sup>89</sup> | 1996 | 133 | Olanzapine | Yes | Yes | Yes |
| Borison 1996<sup>62</sup> | 1996 | 55 | Placebo | Yes | Yes | Yes |
| Borison 1996<sup>62</sup> | 1996 | 54 | Quetiapine | Yes | Yes | Yes |
| van Kammen 1996<sup>51</sup> | 1996 | 105 | Sertindole | Yes | Yes | Yes |
| van Kammen 1996<sup>51</sup> | 1996 | 48 | Placebo | Yes | Yes | Yes |
| Zborowski 1995a<sup>55</sup> | 1995 | 116 | Placebo | Yes | Yes | Yes |
| Zborowski 1995a<sup>55</sup> | 1995 | 115 | Haloperidol | Yes | Yes | Yes |
| Zborowski 1995b<sup>55</sup> | 1995 | 117 | Sertindole | Yes | Yes | Yes |
| Clark 1972a<sup>43</sup> | 1972 | 19 | Chlorpromazine | Yes | No | No |
| Clark 1972a<sup>43</sup> | 1972 | 18 | Placebo | Yes | No | No |
| Clark 1972b<sup>43</sup> | 1972 | 18 | Loxapine | Yes | No | No |
| Clark 1970a<sup>61</sup> | 1970 | 15 | Chlorpromazine | Yes | No | No |
| Clark 1970a<sup>61</sup> | 1970 | 14 | Placebo | Yes | No | No |

